# Globalized low-income countries may experience higher COVID-19 mortality rates

**DOI:** 10.1101/2020.03.31.20049122

**Authors:** Rodolfo Jaffé, Mabel Ortiz, Klaus Jaffé

## Abstract

Understanding the factors underpinning COVID-19 infection and mortality rates is essential in order to implement actions that help mitigate the current pandemic. Here we evaluate how a suit of 15 climatic and socio-economic variables influence COVID-19 exponential growth-phase infection and mortality rates across 36 countries. We found that imports of goods and services, international tourism and the number of published scientific papers are good predictors of COVID-19 infection rates, indicating that more globalized countries may have experienced multiple and recurrent introductions of the virus. However, high-income countries showed lower mortality rates, suggesting that the consequences of the current pandemic will be worse for globalized low-income countries. International aid agencies could use this information to help mitigate the consequences of the current pandemic in the most vulnerable countries.

## Introduction

Understanding the factors underpinning COVID-19 infection and mortality rates is essential in order to implement actions that help mitigate the current pandemic. SARS-CoV-2 (the virus causing COVID-19 disease) might exhibit a preference for cool and dry conditions, like its SARS-CoV predecessor (Araujo & Naimi, 2020). For instance, recent analyses found an association between temperature and COVID-19 growth rates (Shi et al., 2020; Ficetola & Rubolini, 2020), although another study reported no effect of climate on COVID-19 transmission rates (Luo et al., 2020). Detailed models assessing how climatic and socio-economic factors influence both infection and mortality rates based on sufficient cross-country exponential growth-phase data are nevertheless still lacking.

Here we evaluate how a suit of 15 climate and socio-economic variables influence exponential growth-phase COVID-19 infection and mortality rates across countries with sufficient available data. We hope these results help raise awareness of the dear consequences COVID-19 may impose on some low-income countries.

## Methods

### Epidemiological data

Country-level data on COVID-19 confirmed number of cases and deaths was downloaded from the European Centre for Disease Prevention and Control (https://www.ecdc.europa.eu/en/publications-data/download-todays-data-geographic-distribution-covid-19-cases-worldwide), and included all records up to March 30^th^ 2020. To assess COVID-19 growth rate (hereafter infection rate) and mortality rate, we excluded all records before a country reached a minimum threshold of 100 confirmed cases, as well as countries with less than 10 days of records and less than a total number of 1000 confirmed cases. Additionally, we excluded post-exponential growth records in countries where we observed that growth curves were beginning to flatten (China, South_Korea, Iran, Italy, Spain, United States of America, and Austria; exclusion dates are shown in Script_S1). These stringent filtering criteria ensured we had at least 10 continuous days with confirmed records during the exponential growth-phase, for countries that tested a minimum number of 1000 persons.

### Climate and socio-economic data

We gathered country-level climate and socio-economic data using the countries Alpha-2 and Alpha-3 codes and their centroid coordinates (https://gist.github.com/tadast/8827699#file-countries_codes_and_coordinates-csv). Climate data included mean monthly temperature, precipitation and water vapor pressure (calculated between January and March), and was extracted from WorldClim 10 Arc-minutes rasters (https://worldclim.org/data/worldclim21.html) using each country’s centroid. Socio-economic data included GDP per capita (current US$), electric power consumption (kWh per capita), Human Capital Index (HCI), total population, rural population (% of total population), population ages 0-14 (% of total population), population ages 65 and above (% of total population), imports of goods and services (current US$), international tourism (number of arrivals), and scientific and technical journal articles (number), downloaded from the World Bank (https://data.worldbank.org/indicator) and Domestic General Government Health Expenditure (GGHE-D) per Capita (US$), downloaded from the World Health Organization (https://apps.who.int/nha/database/).

### Statistical analyses

To assess country-level COVID-19 infection rates we fitted generalized linear models with a negative binomial distribution of errors (and a logarithmic link function) to account for overdispersion, using the *glm*.*nb* function from the MASS R package (Venables & Ripley, 2002). The daily number of confirmed cases was used as the response variable and date as the predictor. The model’s coefficient for date was then used as a proxy for infection rate during the exponential growth-phase. Mortality rates for each country were calculated as (Total number of deaths / Total number of cases) × 100. We then excluded observations containing missing data (final sample size was 36 countries) and ran linear multiple regressions with either infection rate or mortality rate as response variables, and all possible combinations of up to three non-correlated predictor variables to avoid overfitting. All climate and socio-economic variables plus latitude were included as predictor variables. Models where constructed using the *dredge* function from the MuMIn package (Bartoń, 2019) and a custom script (https://github.com/rojaff/dredge_mc). We then selected the set of best-fitting models using the Akaike Information Criterion (Δ AIC ≤ 2) and used them to calculate confidence Intervals (CI) for model-averaged coefficients and sum of Akaike weights. All analyses were performed in R (see Script_S1).

## Results

Generalized linear models with a negative binomial distribution of errors described well most COVID-19 growth curves from our final dataset of 36 countries (Fig. 1). We found substantial variation in infection rates (glm coefficients) across countries, which ranged between 1.22⨯10^−07^ (Denmark) and 3.29⨯10^−06^ (United States; Fig. 1). Mortality rates ranged between 0.17 (South Africa) and 9.25 (Indonesia; Fig. 2).

**Figure 1:**
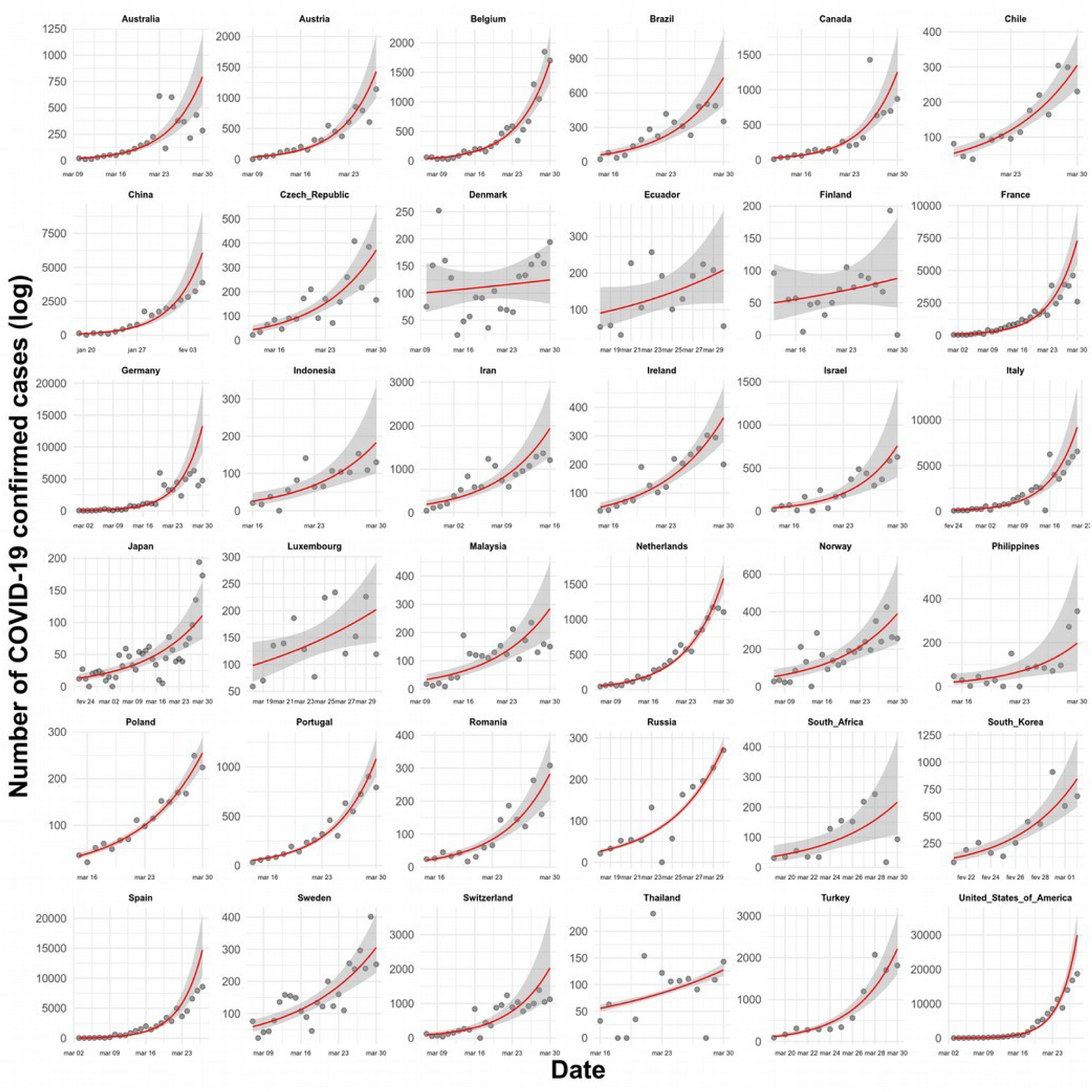
Number of COVID-19 confirmed cases through time in 36 countries. Red lines represent fitted values from generalized linear models with a negative binomial distribution of errors (and a logarithmic link function), and standard errors are shown as shaded areas. Note that post-exponential growth records were excluded in countries where we observed that growth curves were beginning to flatten (see methods for details).

**Figure 2:**
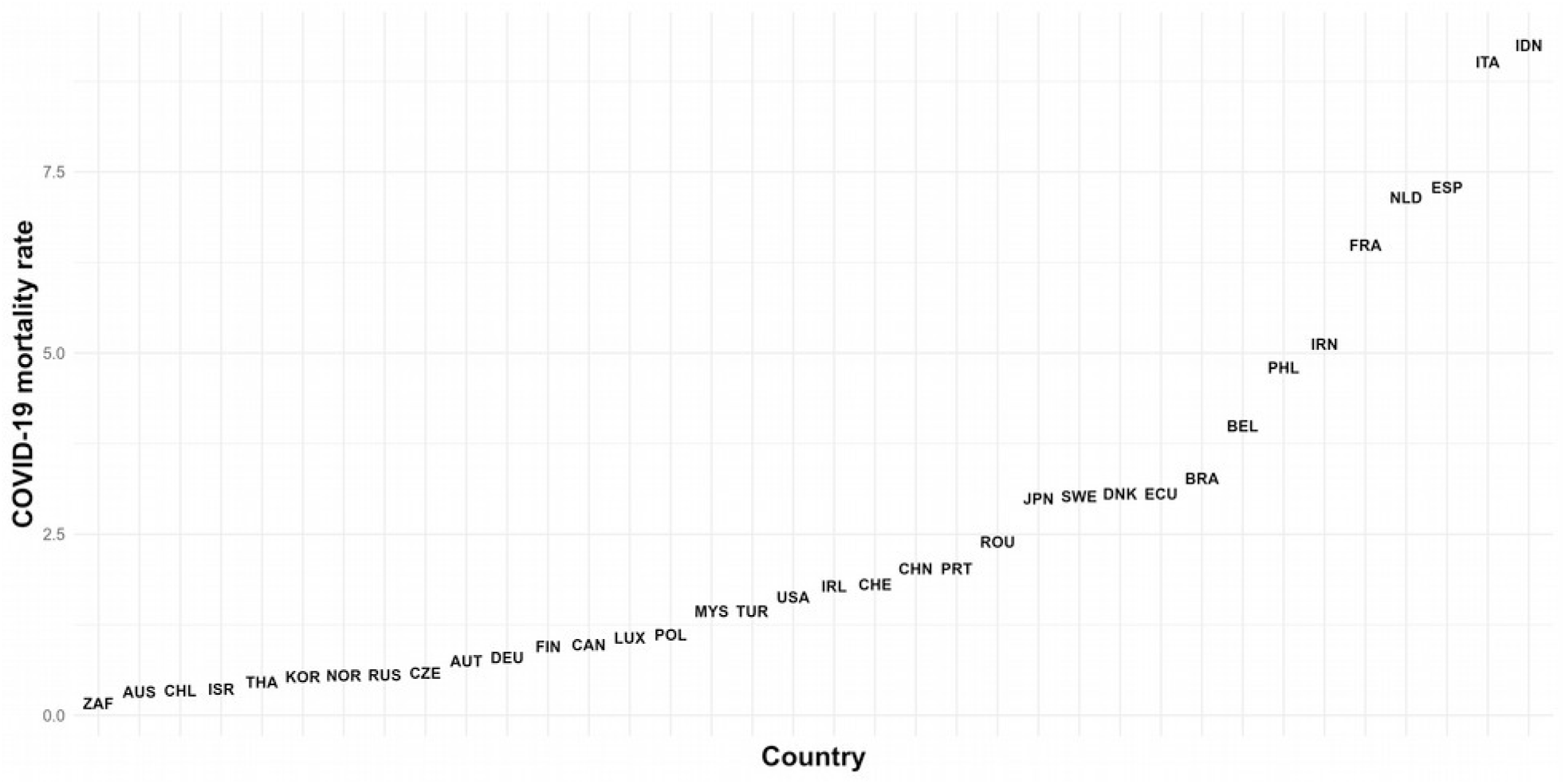
COVID-19 mortality rates across 36 countries. Data points are represented by country Alpha-3 codes (see full legend in Dataset_S1 or here: https://gist.github.com/tadast/8827699#file-countries_codes_and_coordinates-csv).

From the 15 predictor variables assessed, only three were found to be significantly associated with COVID-19 infection rates and two with mortality rates (Table 1). Infection rates were associated with imports of goods and services, international tourism, and published scientific articles (Fig. 3), and were not affected by climatic variables (Table 1). Likewise, local climate did not alter mortality rates, which were most strongly associated with electric power consumption and population of age 65 and above (Fig. 4).

**Table 1:** Confidence Intervals (CI) for model-averaged coefficients and sum of Akaike weights considering the set of best-fitting models (Δ AIC ≤ 2). All models where linear multiple regressions containing either infection rate or mortality rate as response variables (see methods for details). Predictor variables in bold fonts represent those where CI do not contain zero.

| Response | Predictor | Lower CI (2.5%) | Upper CI (97.5%) | Sum of weights |
| --- | --- | --- | --- | --- |
| Infection rate | **log(Articles)** | $1.88 \times 10^{-07}$ | $5.15 \times 10^{-07}$ | 0.37 |
| | **log(Tourists)** | $2.39 \times 10^{-07}$ | $6.49 \times 10^{-07}$ | 0.34 |
| | Population_65+ | $-9.01 \times 10^{-08}$ | $7.18 \times 10^{-11}$ | 0.3 |
| | **log(Imports)** | $2.25 \times 10^{-07}$ | $6.68 \times 10^{-07}$ | 0.23 |
| | log(Water vapor pressure) | $-5.46 \times 10^{-07}$ | $8.26 \times 10^{-09}$ | 0.19 |
| | Government health expenditure | $-2.61 \times 10^{-10}$ | $-2.63 \times 10^{-11}$ | 0.17 |
| | Temperature | $-3.30 \times 10^{-08}$ | $5.15 \times 10^{-09}$ | 0.12 |
| Mortality rate | **log(Electric power consumption)** | -4.27 | -1.20 | 0.76 |
| | **Population_65+** | $2.42 \times 10^{-02}$ | $3.57 \times 10^{-01}$ | 0.39 |
| | log(Imports) | $-2.34 \times 10^{-01}$ | 1.39 | 0.19 |
| | Population_Rural | $-1.40 \times 10^{-01}$ | $4.61 \times 10^{-03}$ | 0.17 |

**Figure 3:**
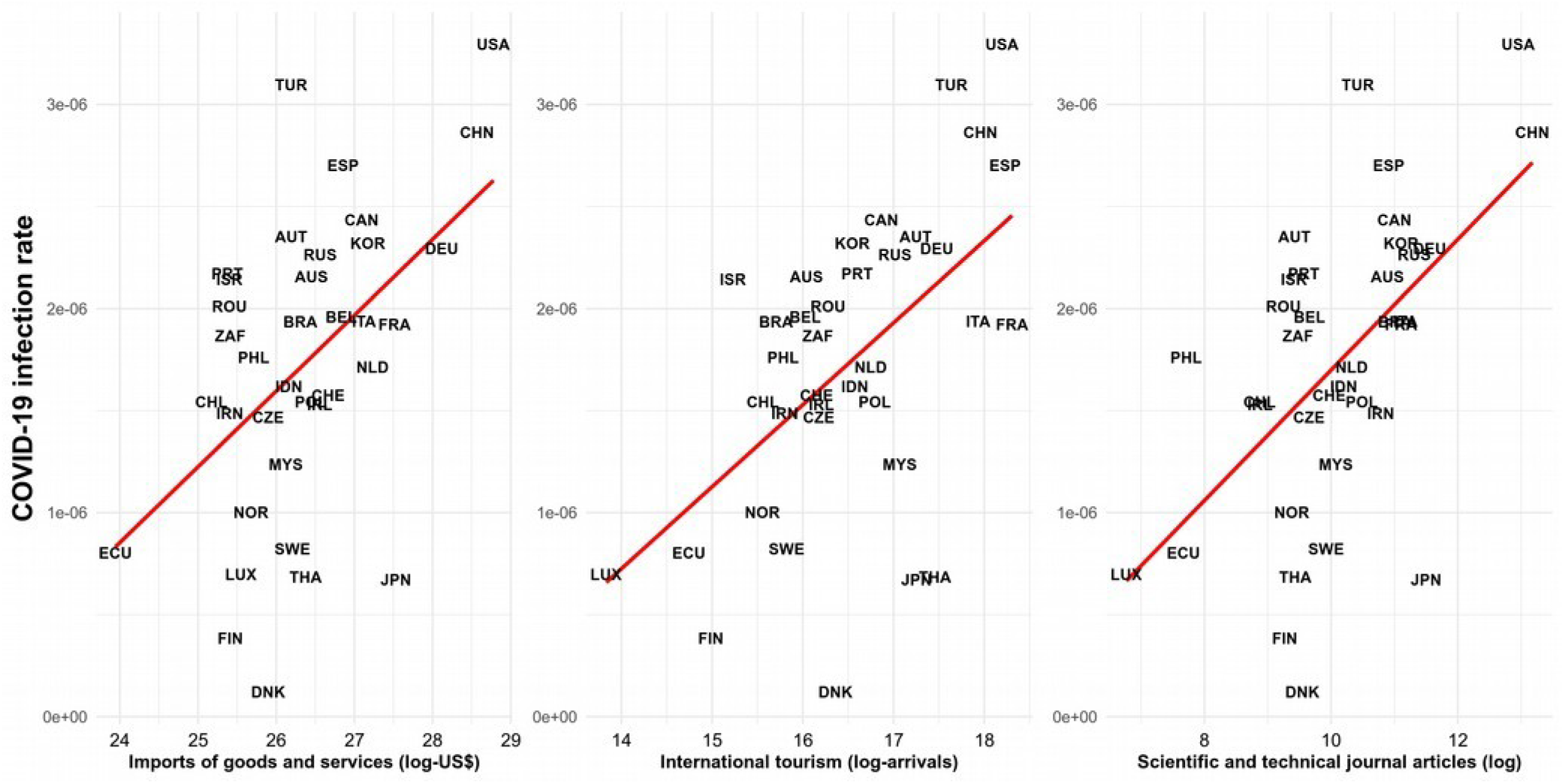
COVID-19 infection rate against imports, international tourism, and scientific articles. All three predictor variables were found among the set of best-fitting models and their model-averaged coefficients did not contain zero. Data points are represented by country Alpha-3 codes (see full legend in Dataset_S1 or here: https://gist.github.com/tadast/8827699#file-countries_codes_and_coordinates-csv).

**Figure 4:**
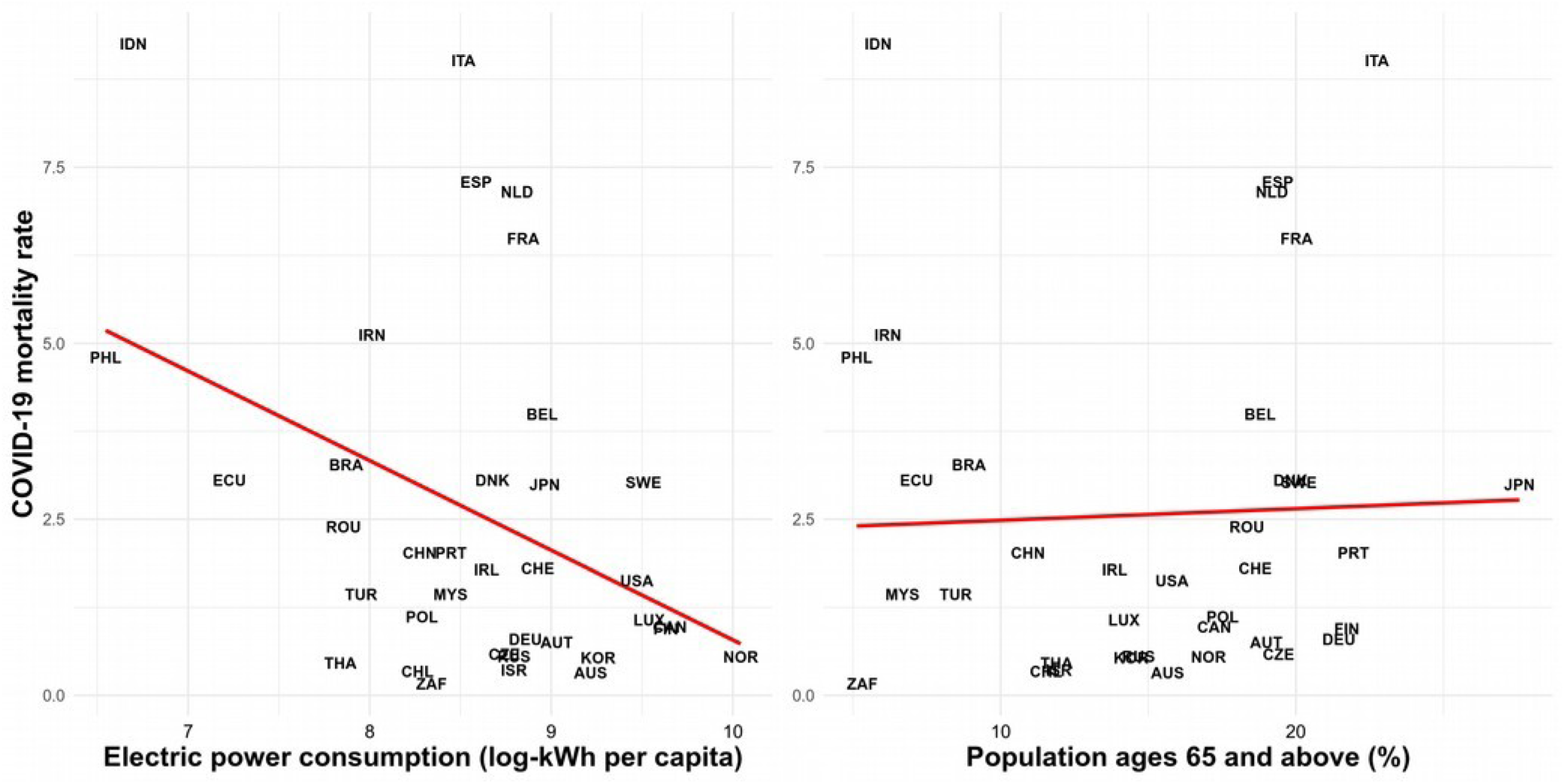
COVID-19 mortality rate against electric power consumption and population of age 65 and above. Both predictor variables were found among the set of best-fitting models and their model-averaged coefficients did not contain zero. Data points are represented by country Alpha-3 codes (see full legend in Dataset_S1 or here: https://gist.github.com/tadast/8827699#file-countries_codes_and_coordinates-csv).

## Discussion

Our analyses suggest that COVID-19 infection rates are affected by imports, international tourism, and scientific productivity (Fig. 3). Since imports of goods and services, international tourism, and the number of scientific articles were highly correlated (Fig. 5), these three variables seem to be representing how open, globalized or integrated countries are (Jaffe et al., 2013). Our findings thus indicate that globalized countries could have experienced multiple and recurrent introductions of the virus via imported goods, tourists, or international exchanges of students or academics (Anzai et al., 2020; Chinazzi et al., 2020). Importantly, local climate (represented by mean temperature, precipitation and vapor pressure) was not found associated with infection rates, supporting a recent study (Luo et al., 2020) and contradicting an evaluation from 31 provincial-level regions in mainland China (Shi et al., 2020) and a global analysis (Ficetola & Rubolini, 2020). The discrepancy between our results and those from Ficetola and Rubolini (2020) may be due to the fact that they calculated exponential growth rates over a period of five days, and assessed a more limited number of predictor variables. Given that we explicitly included the same climatic variables (retrieved from the same source using their described method), and used a larger and more stringent dataset (at least 10 continuous days with confirmed records during the exponential growth-phase, for countries that tested at least 1000 persons), we believe that our statistical analyses should be robust enough to detect an effect of climate should there be one. Our results thus suggest that socio-economic indicators and not climate are influencing COVID-19 infection rates across the 36 analyzed countries.

**Figure 5:**
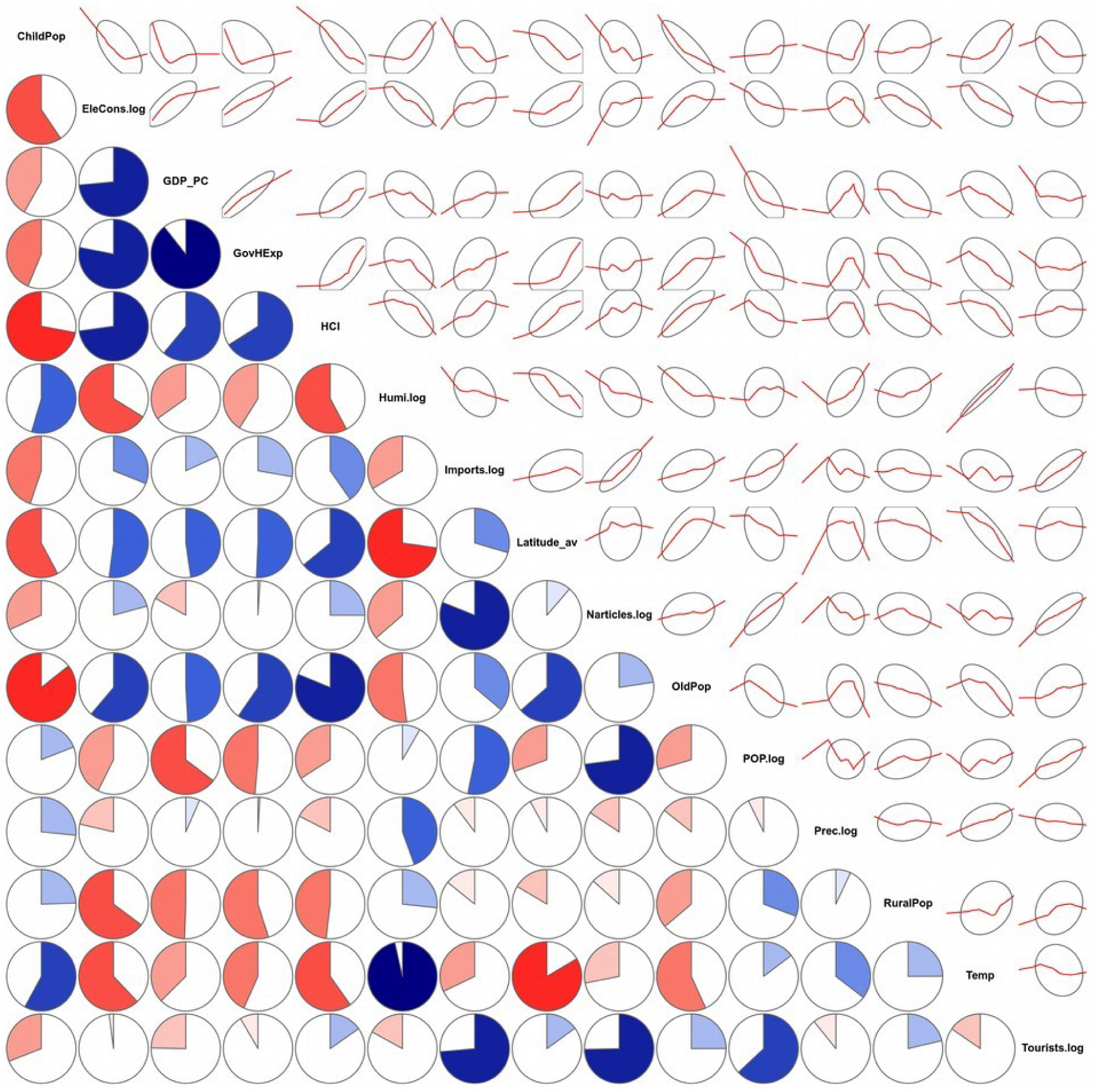
Correlations between all 15 predictor variables. Lower panels show the magnitude and direction of Pearson’s correlation coefficients (blue indicating positive and red negative correlations), while upper panels show a LOESS fit of the data and ellipses encircling most data points. Importantly, electric power consumption, GDP per capita, Government health expenditure and Human Capital Index were highly correlated. Imports, international tourism, and the number of scientific articles were also correlated.

Mortality rates were most strongly associated with electric power consumption and population of age 65 and above (Fig. 4). It is not surprising to find higher mortality rates in countries with a larger population of older people, since this was already identified as the most susceptible population (Chen et al., 2020), and a recent analysis also revealed higher mortality in countries with an older population (Dowd et al., 2020). On the other hand, countries with higher electric power consumption showed lower mortality rates. Since electric power consumption, GDP per capita, government health expenditure, and Human Capital Index were highly correlated (Fig. 5), these indicators seem to be proxies for country-level wealth and development (Narayan & Prasad, 2008; Jaffe, 2013). Our findings thus imply that high-income countries are better placed to face this pandemic, probably due to a higher availability of running water and soap, hospital beds, qualified medical personnel and/or technical equipment. In contrast, low-income countries seem more vulnerable to COVID-19 since they are likely to experience higher mortality rates than countries with better and more accessible services.

## Conclusions

Our study shows important variation in infection and mortality rates across countries, which was primarily explained by socio-economic factors. Importantly, our findings reveal that low-income country receiving large numbers of imported goods and international visitors are likely to exhibit higher COVID-19 infection and mortality rates. International aid agencies could use this information to help mitigate the consequences of the current pandemic in the most vulnerable countries (Bedoya & Dolinger, 2020).

## Data Availability

All data were retrieved from open-access repositories and links are provided in the text.

https://www.ecdc.europa.eu/en/publications-data/download-todays-data-geographic-distribution-covid-19-cases-worldwide

https://gist.github.com/tadast/8827699#file-countries_codes_and_coordinates-csv

https://worldclim.org/data/worldclim21.html

https://data.worldbank.org/indicator

https://apps.who.int/nha/database/

